# Quantifying the impact of a broadly protective sarbecovirus vaccine in a future SARS-X pandemic

**DOI:** 10.1101/2024.08.12.24311730

**Authors:** Charles Whittaker, Gregory Barnsley, Daniela Olivera Mesa, Daniel J Laydon, Chee Wah Tan, Feng Zhu, Rob Johnson, Patrick Doohan, Gemma Nedjati-Gilani, Peter Winskill, Alexandra B. Hogan, Arminder Deol, Christinah Mukandavire, Katharina Hauck, David Chien Boon Lye, Lin-Fa Wang, Oliver J. Watson, Azra C Ghani

**Affiliations:** MRC Centre for Global Infectious Disease Analysis & Abdul Latif Jameel Institute for Disease and Emergency Analytics, School of Public Health, Imperial College London, London, UK; London School of Hygiene and Tropical Medicine, London, UK; Programme in Emerging Infectious Diseases, Duke-NUS Medical School, Singapore; Infectious Diseases Translational Research Programme, Yong Loo Lin School of Medicine, National University of Singapore, Singapore; School of Population Health, Faculty of Medicine and Health, University of New South Wales, Sydney, New South Wales, Australia; Coalition for Epidemics Preparedness Innovations, Oslo, Norway; National Centre for Infectious Diseases, Singapore; Department of Infectious Diseases, Tan Tock Seng Hospital, Singapore; Lee Kong Chian School of Medicine, Nanyang Technological University, Singapore; Yong Loo Lin School of Medicine, National University of Singapore, Singapore; SingHealth Duke-NUS Global Health Institute, Singapore

## Abstract

COVID-19 has underscored the need for more timely access to vaccines during future pandemics. This has motivated development of broad-spectrum vaccines providing protection against viral families, which could be stockpiled ahead of an outbreak and deployed rapidly following detection. We use mathematical modelling to evaluate the utility of a broadly protective sarbecovirus vaccine (BPSV) during a hypothetical SARS-X outbreak, including ring-vaccination, spatial targeting and mass vaccination of high-risk populations. Our results show BPSV ring- or spatially-targeted vaccination strategies are unlikely to contain a SARS-CoV-2-like virus but could contain or slow the spread of a SARS-CoV-1-like virus. Vaccination of high-risk populations with the BPSV ahead of a virus-specific vaccine (VSV) becoming available could substantially reduce mortality. For a 250-day VSV development timeline, BPSV availability reduced infection-related deaths in our model by 54% on average, though exact impact depended on the non-pharmaceutical intervention (NPI) scenario considered. We further show that BPSV availability enables shorter and less stringent NPIs to be imposed whilst limiting disease burden to that observed in the VSV-only scenario, though results are sensitive to vaccine properties (e.g. efficacy), health system capabilities (e.g. vaccination rollout speed) and the assumed timeline to VSV availability. Our modelling suggests that availability of a BPSV for those aged 60+ years could have averted 40-65% of COVID-19 deaths during the pandemic’s first year, with exact impact depending on the size of the maintained stockpile. Our work highlights significant potential impact of a BPSV, but that achieving this depends on investment into health systems enabling rapid and equitable access during future SARS-X pandemics.

## Introduction

COVID-19 highlighted the crucial role of vaccination in reducing disease burden and mitigating socio-economic impact during pandemics. An estimated 14-20 million deaths were averted due to vaccinations in their first year of use (*1*) and enabled lifting of societal restrictions that carried significant socio-economic costs (*2*). Development and authorisation of highly efficacious vaccines against COVID-19 within a year represents a significant achievement, given that development pipelines typically take ten or more years (*3*). Despite this accelerated timeline, more than 1.5 million confirmed COVID-19 deaths occurred during this time-period (*4*). Moreover, access to doses was highly unequal between the global north and global south (*5*). This is despite the additional lives that equitable allocation strategies could have saved (*6–9*).

SARS-CoV-2 is unlikely to be the last pandemic faced by the world (*10*). The frequency of pathogen spillover and intensity of consequent epidemics are projected to increase in the future (*11, 12*). This has motivated interest in reducing vaccine development timelines, including recent initiatives aiming to enable development, authorisation and manufacture of vaccines against a novel pathogen within 100 days of identification (*13–15*). Estimates suggest that COVID-19 vaccine availability within this timeframe could have averted almost 10 million deaths, primarily in lower and lower-middle income countries where availability was most delayed (*16*). However, other work has indicated that existing approaches to vaccine development are unlikely to achieve development timelines of less than 250 days (*17*). A further limitation of these approaches is their reactive nature: pathogen-specific vaccine development depends on detecting and sequencing the novel pathogen’s genome. This limits the timeliness of strategies aiming to develop vaccines in response to emergence of a novel pathogen; such delays lead to substantial human mortality and/or the necessity of significant (and costly) control measures in the form of non-pharmaceutical interventions (NPIs).

Research has therefore focussed on alternative approaches to vaccine development that might facilitate more rapid availability. Broad-spectrum vaccines providing protection against multiple viruses in the same family or sub-family could be manufactured and stockpiled ahead of a pandemic, ready for rapid deployment following identification of a novel pathogen outbreak. Several vaccines aimed at providing broad and robust protection to a range of coronaviruses are under development (*18, 19*). Many have demonstrated an ability to induce broad neutralising antibodies in mice; several have demonstrated this in non-human primates. Potent pan-sarbecovirus neutralising antibodies have been identified in humans previously infected by a range of coronaviruses (*20–22*), suggesting that vaccines eliciting broad-spectrum protection should be possible. These candidates span a range of approaches, including mosaic nanoparticles containing spike receptor binding domains from multiple sarbecoviruses (*23*); chimeric spike mRNA vaccines (*24*); and antigens based on epitopes conserved across multiple coronaviruses (*25, 26*).

Here, we use a mathematical modelling framework to evaluate the utility of a broadly protective sarbecovirus vaccine (BPSV) during a hypothetical future sarbecovirus (“SARS-X”) pandemic. We explore implementation strategies including ring- and spatial-vaccination for outbreak suppression, and mass vaccination of vulnerable populations for disease burden mitigation. Our work highlights substantial potential public-health impact from widespread and rapid access to a BPSV during a SARS-X pandemic. However, realising the maximum potential benefit of these novel tools will require investment into diagnostics, surveillance and broader public-health response capabilities (*27, 28*). In doing so, we demonstrate the potential utility of broad-spectrum vaccines as tools to support future pandemic preparedness and response efforts.

## Results

### BPSV ring-vaccination could support outbreak containment efforts of a sarbecovirus similar to SARS-CoV-1 but not SARS-CoV-2

We developed a stochastic branching-process framework to explore the potential for a BPSV to support containment of a hypothetical SARS-X outbreak via a ring vaccination approach (*29*) **(Fig 1A)**. We considered two “archetype” sarbecoviruses – one similar to SARS-CoV-1 (**Fig 1B**, mean generation time 12 days, 0% presymptomatic transmission, 0% asymptomatic infections) and one similar to SARS-CoV-2 (**Fig 1C**, mean generation time 6.75 days, 35% presymptomatic transmission, 15% asymptomatic infections). We assumed ring-vaccination identified 80% of contacts (*30*), BPSV efficacy of 35% against infection, that breakthrough infections in vaccinated individuals have a 35% reduced infectiousness, and a delay of 2 days between identification of a symptomatic index case and their contacts receiving the vaccine. We also carried out detailed sensitivity analyses (see **Supplementary Table 1** and below).

**Figure 1:**
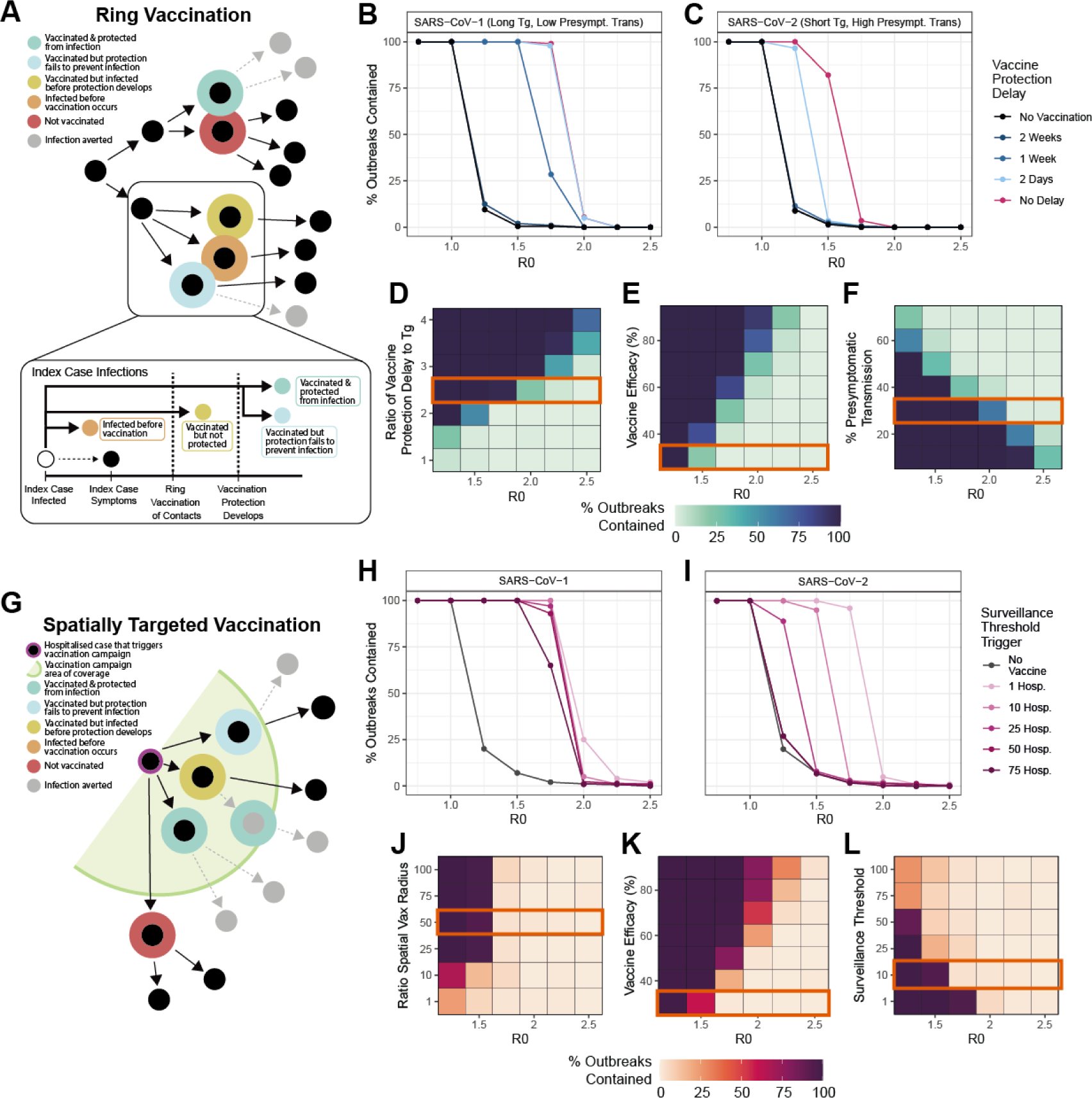
Exploring the prospects for outbreak containment via ring and spatially targeted vaccination strategies using a broadly protective sarbecovirus vaccine (BPSV). A stochastic branching-process-based approach was used to explore the impact of a BPSV on outbreak containment efforts utilising ring or spatially targeted vaccination strategies, and the factors most critical to control. **(A)** Schematic illustrating the ring-vaccination framework. **(B)** % of outbreaks controlled via BPSV ring-vaccination (y-axis) and its dependence on R0 (x-axis), for “SARS-CoV-1-Like” virus. Black line indicates scenario without BPSV, coloured lines indicate different assumptions around vaccine delay to protection (VDP). **(C)** As for **(B)** but for a “SARS-CoV-2-like” virus. **(D)** Sensitivity analysis exploring how the % of outbreaks contained varies with R0 (x-axis) and the ratio of the generation time to the VDP (Tg/VDP). Orange rectangle indicates the value held constant for other sensitivity analyses. **(E)** As for **(D)** but for vaccine efficacy against infection. **(F)** As for **(D)** but % of presymptomatic transmission. **(G)** Schematic illustrating the spatially targeted vaccination framework. **(H)** % of outbreaks controlled (y-axis) and its dependence on R0 (x-axis), for “SARS-CoV-1-like” virus. Black line indicates scenario without BPSV, coloured lines indicate different assumptions around the number of hospitalised cases required to trigger BPSV campaign. **(I)** As for **(H)** but for a “SARS-CoV-2-like” virus. **(J)** % of outbreaks contained by R0 (x-axis) and the ratio of the vaccination campaign spatial radius to the average distance between infections. **(K)** As for **(J)** but for vaccine efficacy against infection. **(L)** As for **(J)** but for surveillance threshold.

Across both pathogen archetypes, the proportion of outbreaks contained decreased as R0 increased and decreases with the delay between vaccination and protection (vaccine delay to protection, VDP). For the “SARS-CoV-1-like-virus”, a VDP of ≤1 week contained all outbreaks for R0≤1.5. However, a VDP of 2 weeks resulted in minimal impact, with less than 1% of outbreaks contained across all values of R0. For the “SARS-CoV-2-like-virus”, with high pre-symptomatic transmission, containment was only possible for VDP≤2 days and R0≤1.5. Prospects for control are sensitive to vaccine characteristics and pathogen properties **(Fig 1D, 1E & 1F)**. When the VDP equalled or exceeded the mean generation time (Tg/VDP = 1, **Fig 1D**), BPSV ring-vaccination did not contain the outbreak **(Fig 1D)**. However, for relatively longer generation times (Tg/VDP ≥3) containment was achieved in all but the highest R0 scenarios. Increasing vaccine efficacy against infection led to an increasing fraction of successfully contained outbreaks for R0≤2 but failed for higher R0 **(Fig 1E)**. For viruses with higher levels of presymptomatic transmission, containment was less likely **(Fig 1F)**.

### Spatially-targeted vaccination strategies using a BPSV must be accompanied by highly sensitive surveillance systems to be impactful

We explored the impact of spatially-targeted vaccination strategies utilising the BPSV for containment. We assumed that BPSV vaccination would be triggered by detection of a cluster of hospitalised cases. Following detection, and a 2-day operational delay, all individuals within a given spatial radius of the home address of the hospitalised case(s) would be vaccinated **(Fig 1G)**. We used the same pathogen archetypes described above, where 95% of “SARS-CoV-1-like-virus” are infections hospitalised and 5% “SARS-CoV-2-like-virus” infections are hospitalised.

For “SARS-CoV-1-like-virus” with R0<2, spatially-targeted vaccination could contain outbreaks across all surveillance system sensitivities considered **(Fig 1H)**. However, for “SARS-CoV-2-like-virus” **(Fig 1I)**, containment with spatially-targeted vaccination with R0<2 occurred only in highly sensitive surveillance scenarios. We undertook sensitivity analyses examining how the proportion of simulated outbreaks contained varied as a function of: i) R0; ii) the ratio of the vaccination campaign’s radius to the average distance separating infections **(Fig 1J)**; iii) BPSV efficacy **(Fig 1K)**; and iv) the number of hospitalisations required to trigger the vaccination campaign **(Fig 1L)**. Increased vaccination campaign radius, improved vaccine efficacy and increased surveillance sensitivity were all associated with an increased fraction of outbreaks being contained. However, none of the scenarios contained outbreaks with R0>2.

### Vaccination of high-risk populations with a BPSV during a SARS-X outbreak could significantly reduce mortality and limit need for NPIs

We next adapted a dynamical model of SARS-CoV-2 transmission (*1*) to explore the utility of a stockpiled BPSV in providing rapid protection of high-risk groups (here, assumed to be those aged 60+) to reduce disease burden during a hypothetical SARS-X outbreak in a population with a demography matching the median age-distribution of the World Bank’s Upper Middle-Income Countries (UMICs). In our simulations, pathogen spillover is followed by undetected circulation in the community, with resulting hospitalisations leading to pathogen detection and identification. Virus-specific vaccine (VSV) development is triggered at this time, and takes either 250 or 100 days depending on the scenario considered (representing “realistic” and “ambitious” vaccine development timelines (*13*)). After an assumed delay of 7 days (to allow activation of stockpiles), mass vaccination of the at-risk population with the BPSV begins. Vaccination switches to the VSV once it becomes available, which is distributed to those aged 60+ first (including those received the BPSV, who are boosted with the new vaccine) and then to the rest of the population. For both vaccines, we assume administration is via a single dose regimen.

To explore the benefits of a BPSV, we compared a scenario where both the VSV and BPSV are available to a baseline scenario in which only the VSV is available. In both cases, the VSV is available only after the delay associated with its development, whereas the BPSV is available far sooner. We assume the BPSV has 75% efficacy against severe disease and 35% efficacy against infection whilst the future VSV has vaccine efficacy of 95% against severe disease and 55% against infection. We assume a R0 of 2.5, in-keeping with initial estimates of ancestral SARS-CoV-2 in Wuhan (*31*) (see **Figure S3** for results with R0 values). We illustrate impact with vaccination rates of 3.5% of the country’s population per week, such that the 60+ age-group is vaccinated within 4 weeks **(Fig 2A)** – this corresponds to 1.55 million doses per week in a population of 40 million in the representative demography selected (where 15% of the population are aged 60+). We considered a range of scenarios reflecting differences in the stringency, duration and triggers for NPIs **(Fig 2B)** with NPI days ranging from 0 (no intervention) to 80 for VSV development of 100 days, and 0-175 for VSV development of 250 days.

**Figure 2:**
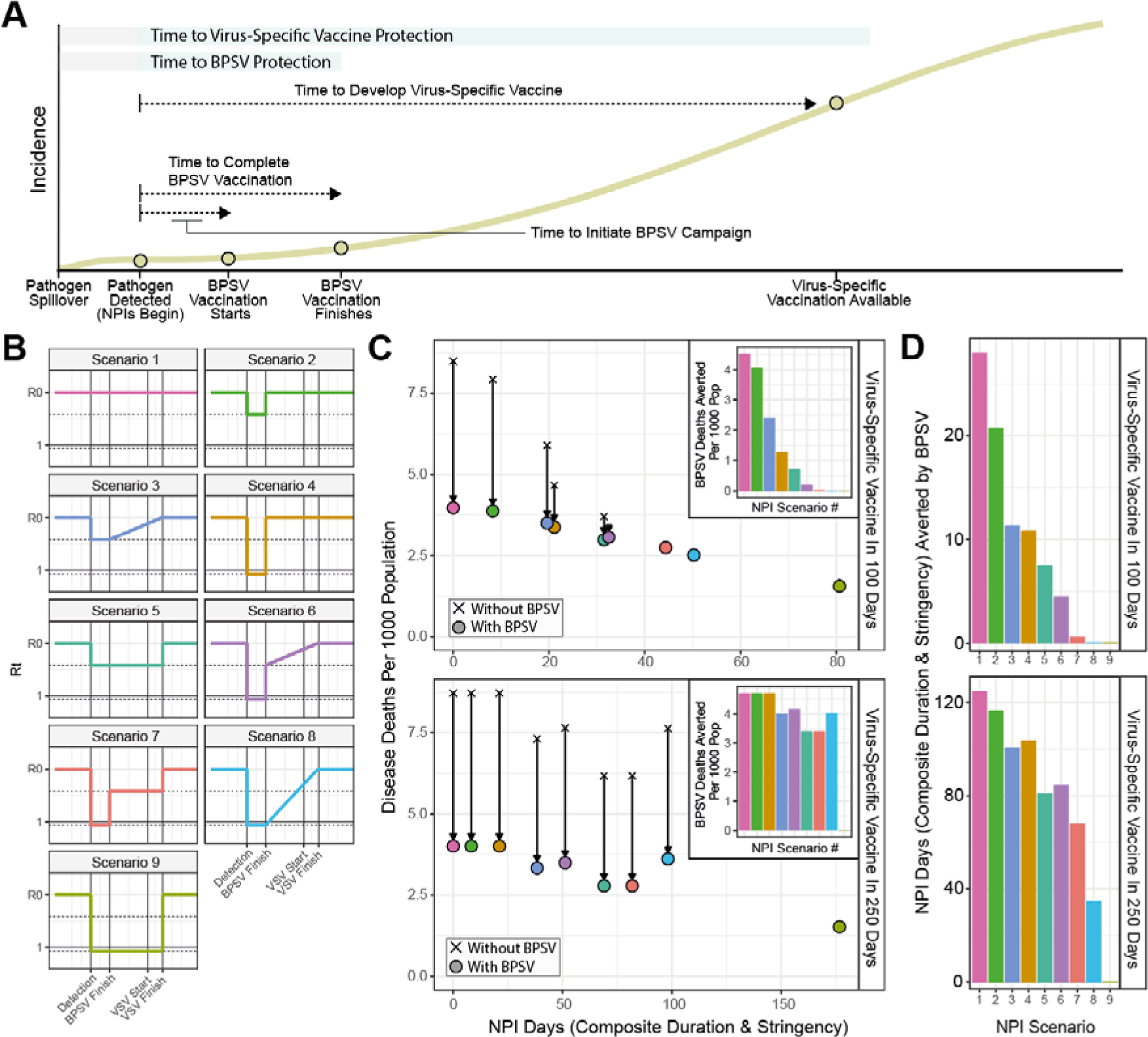
The potential impact of BPSV mass-vaccination campaigns on disease burden during a future SARS-X pandemic. Dynamical modelling of BPSV mass-vaccination of priority groups (those aged 60+) following pathogen detection during a hypothetical SARS-X pandemic. **(A)** Illustrative figure of simulated scenarios and the timing of key events. **(B)** Time-varying reproduction number (Rt) profiles for the different non-pharmaceutical intervention (NPI) scenarios imposed in response to the epidemic that are considered for the analyses presented here. These Scenarios differ by assumed stringency (either no measures, a minimal mandate reducing transmission by 25% or stringent measures reducing Rt to 0.9), duration (either until the BPSV campaign is completed or the disease specific vaccination campaign is completed) and the nature by which these NPIs are relaxed (either instantaneous or gradual). **(C)** BPSV impact on disease burden for each NPI scenario, assuming the VSV is available 100 days (top-panel) or 250 days (bottom-panel) following detection, for an R0 of 2.5. Uncoloured crosses indicate scenario without BPSV (VSV only); points indicate scenarios where BPSV is available, coloured according to NPI scenario. Inset panels shows deaths averted by the BPSV, coloured by NPI scenario. **(D)** BPSV impact on the need for NPIs for the same disease burden. For each NPI Scenario, the Pareto frontier was constructed for the VSV-only scenario, and used to calculate how many fewer NPI days can be imposed in BPSV scenario whilst still limiting limit disease burden to the level observed in the corresponding VSV-only scenario.

The BPSV has the greatest impact if no NPIs are implemented. When the VSV is available after 250 days, projected deaths per 1,000 population were reduced from 8.6 without the BPSV to 3.85 with the BPSV. Similar results are obtained when the VSV is available after 100 days **(Fig 2C, NPI Scenario 1).** With more stringent NPIs, the BPSV averts fewer deaths, but its impact depends on the time taken for the VSV to become available - if the VSV is developed and deployed more rapidly, the relative benefit of the BPSV is reduced. For example, with a short period of stringent NPIs during the BPSV vaccination campaign followed by a minimal set of NPIs afterwards **(Fig 2C, NPI Scenario 6)**, the BPSV reduces deaths from 7.6 to 3.6 per 1,000 population, a 53% reduction if the VSV is available after 250 days. Impact is more limited if the VSV is available after 100 days. More generally, for the 250-day development timeline, availability of the BPSV limits mortality to levels below all but the most stringent NPI scenarios when the BPSV is absent **(Fig 2C, NPI Scenario 9)**, enabling shorter and less stringent NPIs to be in-place for the same total disease burden. BPSV impact was high in R0=3.5 scenarios across all VSV development timelines considered. In R0=1.5 scenarios, BPSV impact was minimal except for the longest VSV development timeline (365 days) (**Fig S3).**

BPSV availability also affects the level of NPIs required to limit disease burden to the level observed when only the VSV is available **(Fig 2D).** For a 250-day development timeline and NPI Scenario 6 (short period of stringent NPIs during BPSV vaccination campaign followed by minimal NPIs), BPSV availability reduced the number of NPI days required from 136 to 51 (63% reduction) **(Fig 2D, lower panel)**. For a 100-day development timeline and the same NPI Scenario, BPSV availability reduced the NPI days by only 4 days (14%) **(Fig 2D, upper panel)**. In general, longer VSV development timelines necessitate more protracted and stringent NPIs to limit accrued disease burden to the same level. This is where BPSV availability of the most reduces NPI need.

### BPSV availability could have substantially reduced mortality during the COVID-19 pandemic

Given the impact that NPIs have on assessing the value of the BPSV, we explored the potential impact that a stockpiled BPSV could have had on COVID-19 mortality in the pandemic’s first year **(Fig 3A)**, using published model fits calibrated to excess mortality data (*16*). We assume BPSV vaccination begins following 1000 globally reported COVID-19 deaths (see **Fig S2** for sensitivity analyses) and with rates of vaccination specific to World Bank income groups (derived from Our World In Data (*32*)). We also varied assumptions about the size of the BPSV stockpile (and hence fraction of eligible population able to be vaccinated) – this ranged from 40% (“Low” coverage) to 80% (“High” coverage) and also explored a scenario in which the size of the stockpile maintained varied by country (“Variable” coverage) in a manner determined by its World Bank Income Group.

**Figure 3:**
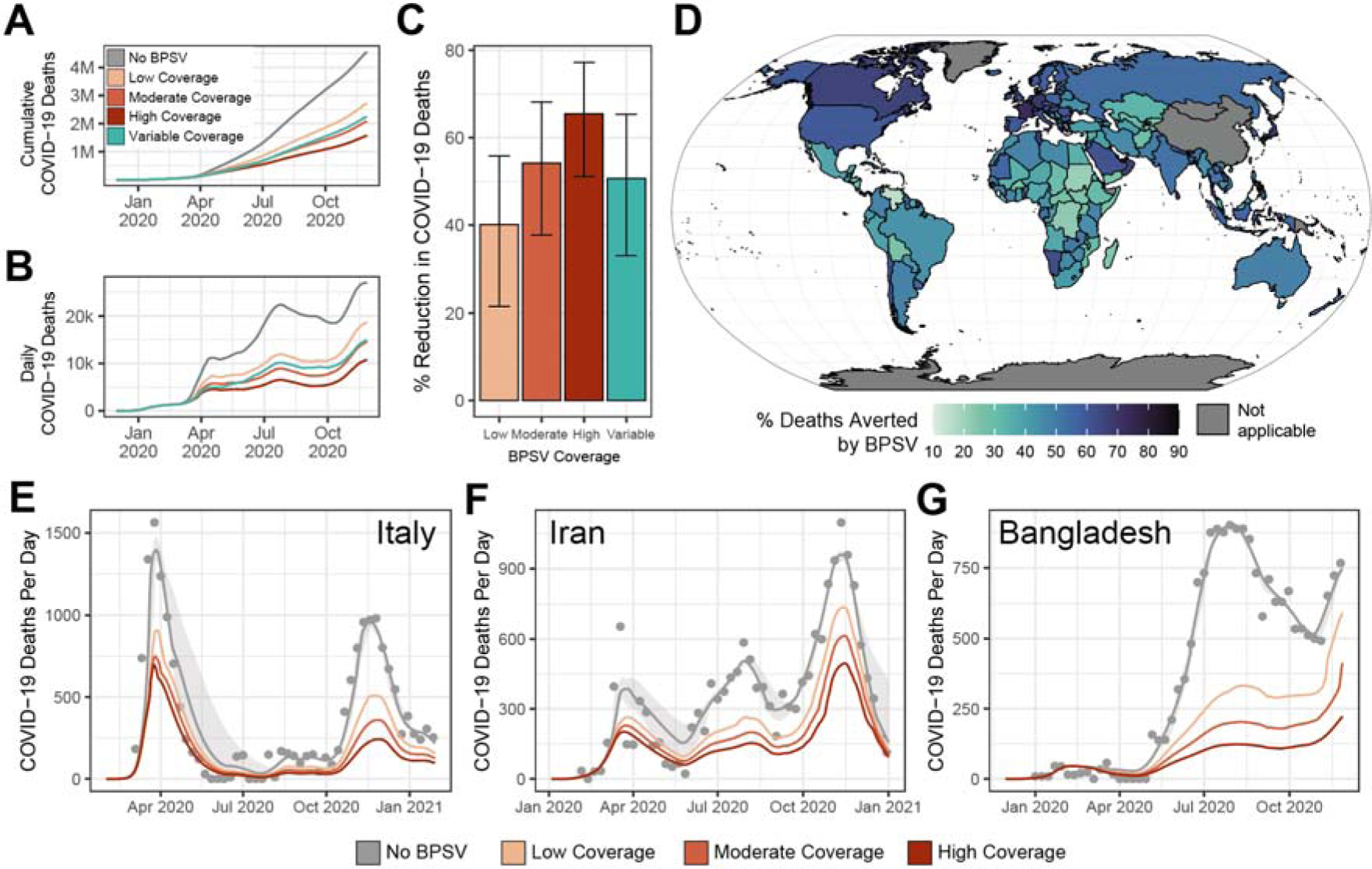
Retrospective evaluation of BPSV impact during the COVID-19 pandemic in selected countries. Analyses using published model fits calibrated to excess mortality data (*16*) retrospectively assessed the potential impact of BPSV availability on COVID-19 mortality during the SARS-CoV-2 pandemic, under various assumptions about the size a BPSV stockpile countries have access to. **(A)** Cumulative global COVID-19 deaths during the first year of the pandemic without (grey) the BPSV, and with the BPSV (coloured lines). Low coverage = BPSV stockpile size sufficient to vaccinate 40% of elderly population; Moderate coverage = 60%; High coverage = 80%. Variable coverage indicates size of stockpile varies according to the World Bank Income Group each country belongs to (LIC = 20%, LMIC = 40%, UMIC = 60%, HIC = 80%). **(B)** As for **(A)** but for daily COVID-19 deaths. **(C)** Modelled impact of the BPSV during the first year of the COVID-19 pandemic in different countries around the world, assuming stockpile size varies by World Bank Income Group (“Variable coverage”). Country colour indicates the percentage of COVID-19 deaths occurring in the first year of the pandemic that could have been averted if a BPSV had been available. **(D)** BPSV impact on COVID-19 mortality in Italy during the first year of its COVID-19 epidemic. Grey line indicates model fit to COVID-19 excess mortality data (light grey points) and ribbon indicates the 95% CI. Coloured line indicates expected mortality when the BPSV is available. Line colours reflects the assumption about the size of the BPSV stockpile. **(E)** As for **(D)** but for Iran instead of Italy. **(F)** As for **(D)**, but for Bangladesh instead of Italy.

Our results indicate that a BPSV stockpile sufficient to vaccinate 60% of the global eligible population could have averted 54% (37%-68%, “Moderate Coverage” scenario) of COVID-19 deaths **(Fig 3A** and **Fig 3B)**, ranging from 40% (21%-56%, “Low Coverage” scenario) to 65% (51%-77%, “High Coverage” scenario) depending on the size of the maintained stockpile **(Fig 3C)**. A global stockpile is unrealistic but could be prioritised in places affected early in the pandemic – assuming nationally maintained stockpiles whose size varied by country averted 50% of deaths (33%-65%, “Variable Coverage” scenario, **Fig 3D**). Under a “Moderate coverage” scenario, in Italy, BPSV availability could have reduced mortality during the first wave from 1360 daily deaths at its peak to 750 deaths **(Fig 3D)** and total COVID-19 mortality over the first year from124,500 to 59,900 deaths, a 52% reduction **(Fig 3D)**. Similar impacts were estimated for the epidemics in Iran **(Fig 3E)** and Bangladesh **(Fig 3F)**. Respective stockpile sizes required for these countries to vaccinate their eligible populations and achieve this impact would have been 10.8 million doses for Italy, 5.2 million doses for Iran and 7.9 million doses for Bangladesh.

### BPSV impact depends on target product characteristics

The presented results make assumptions about the properties of a hypothetical BPSV. The majority of BPSV candidates are at preclinical stages and their potential properties remain uncertain (*18*). We therefore undertook detailed sensitivity analyses to understand how different BPSV properties shape its potential impact, whilst also varying the time the time taken to develop the more efficacious VSV and features of the pathogen (see **Supplementary Table 2** for list of sensitivity analyses). We considered three possible NPI response to the epidemic: i) Minimal (25% reduction in Rt during BPSV campaign, no NPIs thereafter); ii) Moderate (25% reduction in Rt during BPSV campaign, gradual lifting of NPIs until VSV campaign completes); and iii) Stringent (Rt<1 during BPSV campaign, slow cessation of NPIs until VSV campaign completes). Our results show that increasing BPSV severe disease efficacy reduces of mortality in all but the lowest R0 scenarios **(Fig 4A).** For low R0 (R0 = 1.5, **Fig S3**) and VSV development time of 250 days, BPSV impact is minimal under all NPI scenarios and disease efficacy values considered, with <1.5 deaths per 1,000 population averted. Under these scenarios, VSV development is accomplished before significant spread through the population. For higher R0, increasing BPSV efficacy increases averted mortality **(Fig 4A** and **Fig S3A)**. We observed a less marked influence of infection efficacy on BPSV impact (**Fig 4B** and **Fig S3C**). This is because the BPSV campaign only targets a small fraction of the population (those aged 60+) and therefore the impact on onwards transmission is limited. BPSV impact increases with longer duration of elicited immunity **(Fig 4C** and **Fig S3B)**.

**Figure 4:**
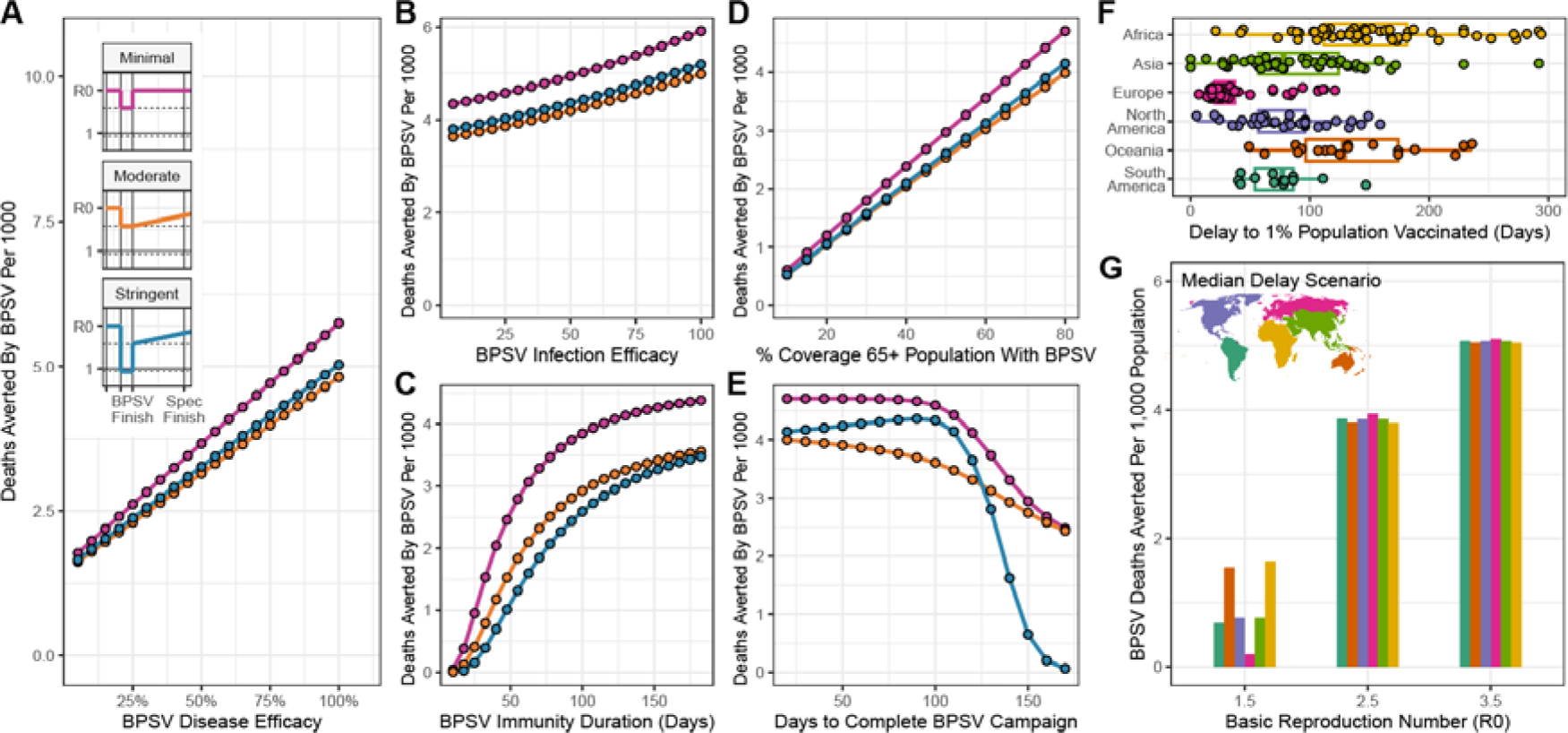
Dependence of BPSV impact on intrinsic vaccine properties and vaccination campaign dynamics. Sensitivity analyses exploring the sensitivity of BPSV impact to intrinsic BPSV properties and factors governing the speed, availability and coverage of the BPSV vaccination campaign. **(A)** Deaths averted by the BPSV (per 1,000 population) and BPSV efficacy against severe disease. Results coloured according to NPI scenario considered (pink = minimal, orange = moderate, blue = stringent), for R0=2.5. Inset panels show Rt profile for each NPI scenario. Assumed virus-specific vaccine (VSV) development timeline was 250 days. **(B)** As for (A) but for BPSV efficacy against infection. **(C)** As for (A) but for BPSV immunity duration. **(D)** As for (A) but for BPSV stockpile size (and associated coverage of the target population that can be achieved). **(E)** As for **(A)**, but for the rate of vaccination during the BPSV campaign (and the associated time taken to vaccinate all eligible and willing individuals). **(F)** The delay (in days) between the first country in the world achieving 1% of their population vaccinated with COVID-19 vaccines and other countries achieving this same milestone. Individual coloured points are specific countries – data from Our World In Data. **(G)** Impact of delays to BPSV access on deaths averted per 1,000 population. Scenarios shown are for moderate NPIs and with continent-specific VSV access delays derived from (F).

### BPSV impact is shaped by stockpile size, vaccination campaign speed and access equity

We additionally carried out analyses exploring how BPSV impact is affected by procurement and health system factors shaping the speed and size of the vaccination campaign. BPSV impact increased linearly with the size of the stockpile **(Fig 4D** & **Fig S3D)**. Assuming an R0 of 2.5, and maintaining a stockpile sufficient to vaccinate 76% of eligible 60+ (in-keeping with estimates of primary SARS-CoV-2 vaccination coverage of older adults as of December 2022 (*33*)), a BPSV could avert 3.1 deaths per 1,000 population. BPSV impact is positively correlated with vaccination campaign speed **(Fig 4E)**. For the moderate NPI scenario, 3.8 deaths per 1,000 are averted when the campaign is completed in <2 months, versus 2.7 deaths per 1,000 for a 5-month campaign. However, under the stringent NPI scenario, there was no additional impact of the BPSV if the campaign took more than 5 months. Vaccination campaign speed had even more influence on BPSV impact when R0=3.5, and minimal impact when R0=1.5 **(Fig S3E).**

We also explored the impact of delays to VSV access. During the COVID-19 pandemic, countries in the global south experienced substantial delays before receiving vaccines (*9*), illustrated by the time taken to achieve 1% vaccination coverage (as a proxy for timeliness of vaccine access) (*32, 34*) **(Fig 4F)**. Relative to the earliest country vaccinating 1% of their population, European countries experienced a median delay of 32 days (IQR 27-39), whereas for African countries this was 135 days (IQR 110-180). We incorporated these delays into the VSV development timeline and evaluated BPSV impact. For a VSV development time of 250 days, a moderate NPI scenario and R0 = 3.5, the BPSV averted significant mortality, across all vaccine access delays considered **(Fig 4G)**. However, for R0 = 1.5, the BPSV had a substantially higher impact on disease burden when access was delayed to a level similar to that experienced by the average African (1.75 deaths averted per 1,000 population) than the European delay scenario (0.18 deaths averted per 1,000 population).

### Surveillance system sensitivity and timeliness of virus detection initiation shapes BPSV impact

Initiation of BPSV vaccination and VSV development depend on detection and identification of the novel virus. We carried out sensitivity analyses exploring how surveillance sensitivity affects BPSV impact. Assuming R0 of 2.5, a moderate NPI scenario and VSV development within 100 days, BPSV impact was lowest when surveillance sensitivity was high (i.e. a low hospitalisation threshold for response trigger), and highest when surveillance sensitivity is low (i.e. a higher hospitalisation threshold for response trigger) **(Fig 5B left hand panel)**. Where surveillance sensitivity is high and VSV development is rapid, the VSV can be distributed before significant epidemic progression, and thus there is minimal additional impact of BPSV availability. In contrast, BPSV impact was high under all surveillance sensitivity scenarios considered when VSV development was 250 days **(Fig 5B right hand panel)**. In this context, VSV development occurred after the epidemic and so there was significant benefit to availability of a BPSV and the protection it provided to eligible individuals.

**Figure 5:**
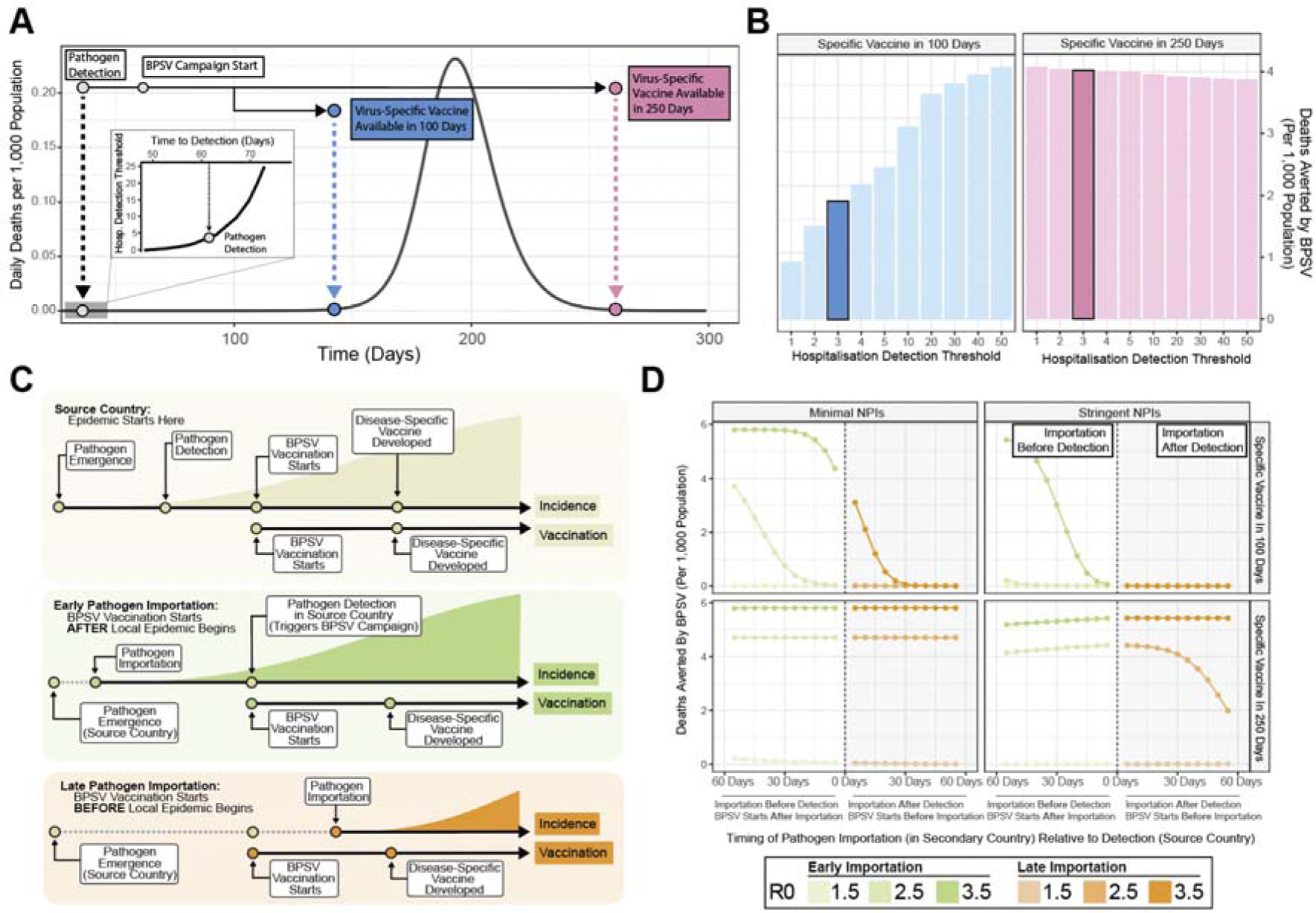
Surveillance system sensitivity, time-to-detection and BPSV impact. Sensitivity analyses exploring the influence of surveillance sensitivity and timing of detection on BPSV impact. **(A)** Epidemic time-series (dark grey line). Inset panel shows time-to-detection (x-axis) versus hospitalisation detection threshold (daily hospitalisations required for initial virus detection to occur). Coloured points and arrows indicate timing of virus-specific vaccine (VSV) developed under timelines of 100 (blue) or 250 (pink) days. **(B)** Deaths averted by BPSV per 1,000 population according to hospitalisation detection threshold and VSV development timeline. Highlighted bars indicate the hospitalisation detection threshold shown in (A). **(C)** Timeline illustrating the distinction between source country (where the epidemic originates) and secondary countries (where epidemics are seeded via importations from the source country). Secondary countries include “early importers” (virus importation occurs before detection in source country); and “late importers” (virus importation occurs after detection in the source country). **(D)** Deaths averted by BPSV (y-axis) against the number of days that virus detection in the source country is ahead of or after pathogen importation (to the secondary country). Grey-shaded regions indicate virus importation occurs after detection in source country. White regions indicate virus importation occurs before detection in source country.

### Timeliness of BPSV campaign initiation relative to the timing of pathogen importation influences impact

The analyses presented thus far have been restricted to understanding BPSV impact in the country where the novel virus outbreak starts. This assumes that BPSV vaccination is initiated in response to pathogen identification following hospitalisations in local facilities. However, both SARS-CoV-1 and SARS-CoV-2 pandemics exhibited early international spread. We carried out analyses exploring how timeliness of novel pathogen detection in the country where the virus initially emerges (“source country”) might influence the impact of public health responses utilising the BPSV in other countries (“secondary countries”). Here, virus detection in the source country is assumed to trigger VSV development and BPSV vaccination in secondary countries. We vary importation timing relative to detection such that importation into the secondary country occurs either before (“early importation”) or after (“late importation”) detection in the source country. In both, we assume BPSV vaccination starts when the virus is detected in the source country **(see Fig 5C)**. Assuming the VSV is available 100 days after detection, the BPSV had limited impact in the late importation scenario because only limited community transmission occurs before the VSV becomes available **(Fig 5D, top row, orange lines)**. In contrast, the BPSV had significant impact in the early importation scenario, especially for high R0 (R0 = 2.5 and 3.5). Under these scenarios, significant community transmission and spread occurs before the VSV becomes available, and the BPSV therefore effectively protects high-risk populations **(Fig 5C, top row, green lines)**. For a VSV development time of 250 days, the BPSV had substantial public health impact under most scenarios considered.

## Discussion

Our research shows that timely BPSV access could reduce both the disease burden and socio-economic impacts of future SARS-X pandemics. Notably, an available BPSV stockpile in the country where the new virus emerges could, with only limited NPIs, significantly lower mortality to below what could be achieved with longer and more stringent NPIs alone. Such a vaccine therefore has the potential to both reduce disease burden and the wider economic costs associated with NPIs. Several factors influence this impact. These act at two levels; those intrinsic to properties of the BPSV (which affect the quality of protection it offers) and those that influence the number of individuals infected between BPSV vaccination finishing and the VSV becoming available. It is during this period that the BPSV provides protection to individuals who would otherwise be infected or hospitalised. Faster development timelines for the VSV, or low infection rates (due to stringent NPIs), can diminish the size of the population infected in this period, reducing the impact of the BPSV campaign. Higher transmissibility (R0), limited NPIs and/or longer VSV development times increase the size of the population infected before the VSV becomes available, increasing the value of a BPSV. The impact of the BPSV is then determined by the size of the maintained BPSV stockpile and the rate at which BPSVs can be delivered to the high-risk populations.

Despite this promise for disease burden reduction, our results indicate that BPSVs are unlikely to greatly enhance early containment, especially for highly transmissible viruses similar to SARS-CoV-2. A critical issue here is the vaccine delay to protection (VDP, the delay between vaccination and protection), with our results suggesting a negligible fraction of outbreaks contained via ring-vaccination when the VDP is of a similar timescale to the generation time of the virus. Given this, other broad-spectrum medical countermeasures such as monoclonal antibodies (where protection arises near-immediately) could represent crucial additions to the arsenal of tools available for outbreak containment (*35*), (*36*), as could tools eliciting more robust immunity against infection and onwards transmission, such as has been observed for vaccines delivered intranasally (*37, 38*). Our results suggest spatially targeted vaccination strategies are unlikely to contain a “SARS-CoV-2-like-virus” except when the spatial area covered by the campaign is large or hospital surveillance is highly sensitive.

Whilst surveillance limitations may hinder BPSV use for containment, its use for disease-burden mitigation is robust to these limitations. This is because surveillance capabilities influence both the timing of the BPSV campaign and the initiation of VSV development. In low-sensitivity systems, which detect pathogens later, both BPSV distribution and VSV development are delayed – this shift in the timing of both events does not significantly change the size of the population infected in the VSV-only scenario (and who are therefore protected by the BPSV in the scenario where it is available). In high-sensitivity systems, early pathogen detection enables prompt BPSV deployment, before significant spread occurs. Previous work has highlighted both the current limitations novel pathogen surveillance capabilities globally (*28, 39*) and the substantial investment still required to address this (*40*). Tools (like the BPSV) that are robust to current limitations in are therefore likely to prove useful.

A significant limitation of the results presented relates to uncertainty around BPSV properties. Multiple candidates are under development (*18*), but no human immunogenicity evaluations have been carried out to date (*23, 41*). This is important given the diversity of coronaviruses that humans are routinely exposed to (*42*) and how past exposure shapes antibody responses to sarbecovirus infection (*43*). To mitigate this limitation, we assumed BPSV efficacy estimates that are lower than those achieved by mRNA vaccines against ancestral SARS-CoV-2 lineages (*44, 45*). We also analysed how BPSV impact depends on vaccine properties such as disease efficacy and immunity duration. Our results show the BPSV has significant impact, especially when VSV development timelines are similar to those achieved with SARS-CoV-2. While the eventual properties of developed BPSVs are uncertain, our results suggest timely BPSV availability and stockpiling can achieve significant public health impact, even when efficacy is lower than disease-specific alternatives. A further limitation is that we do not account for existing cross-immunity to a new SARS-X virus. Previous work in SARS-CoV-1 survivors has shown antibodies providing cross-protection to ancestral and variant SARS-CoV-2 virus (*22*). Given the global prevalence of SARS-CoV-2, future SARS-X burden (and BPSV impact) may be mitigated by cross-immunity. However, this remains highly uncertain and indeed, cross-immunity may serve to further increase the impact of a BPSV or expand the range of possible implementation strategies (e.g. as a booster providing more robust immunity against subsequent variant lineages).

Whilst our work highlights the significant impact of a BPSV, evaluation of the economic viability of maintaining a stockpile is also required. Currently, this is challenging, due to uncertainty around the eventual properties of developed BPSVs and the cost of acquisition, stockpiling and administration. Ring-vaccination strategies for Ebola have been suggested to be cost-effective (*46*), whilst previous research has highlighted COVID-19 vaccinations as consistently cost-effective or cost-saving (*47*). Whilst the outsized economic costs of the COVID-19 pandemic (*2*) suggest that maintenance of a BPSV stockpile would be cost-effective, uncertainty in the timing and scale of future outbreaks alongside the costs associated with maintenance of a stockpile necessitates further assessment of the cost-effectiveness of different implementation strategies and mechanisms for stockpiling.

Despite these limitations, our work highlights the significant impact that could be achieved through development, manufacture and stockpiling of BPSVs to facilitate rapid access in a hypothetical SARS-X pandemic. Such vaccines could provide an effective way to protect high-risk groups during the period between novel pathogen identification and the development of efficacious VSVs. In doing so, BPSV utilisation has scope to avert both significant disease burden and substantial economic losses through relaxing the requirement for stringent NPIs to control transmission. However, our work also shows that realising the benefits of the BPSV is critically dependent on other feature of the health system, necessitating investments into capabilities enabling rapid vaccine distribution if these broad-spectrum medical countermeasures are to most effectively form a part of future pandemic preparedness strategies.

## Methods

### Stochastic Branching Process Modelling Framework

We extended stochastic branching-process modelling frameworks initially developed to explore SARS-CoV-2 control through contact tracing (*48, 49*) and MERS-CoV vaccination (*50*) to simulate different vaccination strategies focussed on outbreak containment (defined as a final outbreak size of <10,000 infected individuals). These were ring-vaccination (where detection of symptomatic cases triggers reactive vaccination of all contacts of that case, **Fig 1A**) and spatially targeted vaccination (where hospitalised cases trigger vaccination of all individuals in a defined geographic area, **Fig 1G**).

For both strategies, we calculate the proportion of outbreaks contained relative to a scenario in which the BPSV is not available whilst varying the pathogen epidemiological properties, the intrinsic properties of the BPSV, and features of the vaccination campaign response. The epidemiological properties that we vary are R0, generation time distribution, extent of pre-symptomatic transmission, proportion of asymptomatic infections and the probability of being hospitalised. We explore two pathogen “archetypes” – the first has properties similar to SARS-1, with a long generation time, limited pre-symptomatic transmission, low proportion of asymptomatic infections and high disease severity. The second is SARS-CoV-2, with a shorter generation time, extensive pre-symptomatic transmission, more asymptomatic infection and low disease severity. For both archetypes, we vary the R0 across a range of values. Furthermore, we assume that past exposure to SARS-CoV-2 does not generate significant immunity to SARS-X. BPSV properties varied across model runs were efficacy against infection, relative infectiousness of breakthrough infections, and the assumed delay between vaccination and immunological protection developing.

In all instances, results are the proportion of outbreaks contained across 100 stochastic simulations, per parameter combination and vaccination strategy considered. Code to reproduce the results is available at https://github.com/mrc-ide/diseaseX_modelling. For further details of the model and parameterisation, see **Supplementary Information.**

### Dynamical Compartmental Modelling Framework

We adapted a published compartmental model of SARS-CoV-2 transmission and vaccination (*1*) to explore the impact of BPSV availability on disease burden during a future hypothetical SARS-X pandemic. The original model is described in (*16, 51, 52*), with details of the extensions added here described below and in the **Supplementary Information**. Briefly, we extended this modelling framework to enable simulation of two vaccines with distinct properties (the BPSV and the Virus Specific Vaccine, VSV). We model two distinct forms of vaccine efficacy: efficacy against infection and efficacy against severe disease in breakthrough infections. The BPSV, available immediately upon detection, is assumed to have an efficacy of 75% against severe disease and 35% against infection. The disease-specific vaccine, developed later, has a more favourable efficacy profile - 95% against severe disease and 55% against infection, with development timelines of either 100 or 250 days. Following spillover, pathogen detection occurs on the first day with ≥5 daily hospitalisations and leads to initiation of both the BPSV vaccination campaign and disease-specific vaccine development. The timing of detection was calculated using the stochastic branching-process framework. The BPSV is used to vaccinate individuals over the age of 65 years. All age-groups except those under 15 are eligible to receive the disease-specific vaccine, with rollout of this vaccine (sufficient to achieve a coverage of 80% of the population) occurring in the oldest age-groups first.

#### Hypothetical SARS-X Pandemic Scenario Modelling

We assume an R0 of 2.5 and generation time of 6.7 days, aligned with estimates for the original Wuhan-1 strain, as well as a severity profile and age-specific IFR similar to SARS-CoV-2 (*53*), adjusted to give an overall population-level IFR of 1%. We assume a demographic age-structure matching the age-distribution of the World Bank Upper Middle-Income Country with the median age and assume no healthcare constraints that limit the ability of hospitalised individuals to access medical care, noting that relaxing this assumption would increase our estimates of BPSV impact. We explore different scenarios varying the duration, stringency, and timing of imposed NPIs. These scenarios are each generated from three levels of NPI stringency: i) none, keeping Rt equal to R0; ii) minimal, reducing Rt by 25%; iii) stringent, reducing Rt to 0.9. Using these three levels of NPI stringency, we construct three scenarios for the purposes of analysis: NPI Scenario 1) minimal NPIs applied briefly after pathogen detection until the end of the BPSV campaign; NPI Scenario 2) moderate NPIs lasting until the BPSV campaign’s end, then relaxed until the completion of the disease-specific vaccine rollout; NPI Scenario 3) stringent NPIs maintained throughout the BPSV campaign, then eased until the end of the disease-specific vaccine rollout. For each NPI scenario, an “NPI index” (representing a composite of the stringency of imposed NPIs and their duration) was calculated. Deaths averted per 1,000 population by the BPSV were estimated by comparing deaths in scenarios with both BPSV and the disease-specific vaccine to scenarios with only the disease-specific vaccine. Potential NPI days averted by availability of the BPSV were calculated as follows: first, we constructed the Pareto frontier across explored NPI scenarios in the case where only the VSV was available. We then calculated the difference in composite NPI days between the scenario in which the BPSV is available and the composite NPI days for the point lying on the Pareto frontier leading to the same number of deaths as in a scenario where only the VSV is available.

#### Retrospective Evaluation of Potential Impact During SARS-CoV-2 Pandemic

Using published Bayesian country-specific model fits to excess mortality data (*1*) we explored the potential impact that a stockpiled BPSV could have had on COVID-19 mortality in the first year of the pandemic. We first sampled 100 draws from the previously estimated posterior distribution of Rt. To then estimate the impact of BPSV, we simulated a counterfactual scenario for each sampled Rt trajectory in which BPSV vaccines were deployed following cumulative globally reported COVID-19 deaths reaching a defined threshold, assuming all countries possess a BPSV stockpile to vaccinate 60% of their eligible population and with rates of BPSV vaccination specific to each World Bank Income Group and determined based on data from Our World In Data (*32*). Deaths averted by the BPSV were calculated by subtracting the estimated COVID-19 deaths from the simulation with the BPSV from the simulation without the BPSV, with the median deaths averted per 1,000 population reported here. See **Supplementary Information** for further details.

## Author Contributions

**Conceptualization:** CW, GB, DOM, OJW & AG

**Methodology:** CW, GB, DJL, OJW & AG

**Investigation:** CW & AG

**Visualization:** CW, DOM & AG

**Funding acquisition:** OJW & AG

**Project administration:** AG

**Supervision:** AG

**Writing – original draft:** CW & AG

**Writing – review & editing:** All Authors

## Data & Materials Availability

The modelling framework, along with all relevant data and code required to reproduce the analyses presented here are freely available in Github repository (https://github.com/mrc-ide/diseaseX_modelling).

## Data Availability

The modelling framework, along with all relevant data and code required to reproduce the analyses presented in this manuscript are freely available in Github repository (https://github.com/mrc-ide/diseaseX_modelling).

## Supplementary Information

### 1. Supplementary Methods: Branching Process Framework

#### 1.1 Overview of Stochastic Branching Process Modelling Framework

We extended a stochastic branching-process modelling framework initially developed to simulate SARS-CoV-2 transmission and control through contact tracing (*48, 49*), and use this framework to explore a number of different BPSV vaccination strategies. We explore this for two pathogen “archetypes” – the first pathogen archetype has properties similar to SARS-CoV-1, and is characterised by a long generation time, a limited degree of pre-symptomatic transmission, low proportion of asymptomatic infections and high disease severity. The second is SARS-CoV-2, characterised by a shorter generation time, high proportion pre-symptomatic transmission, moderate proportion of asymptomatic infections and low disease severity. For both pathogens, we vary the basic reproduction number across a range of values. Below, we describe this framework in general terms, and provide a table describing the exact parameter values used to model different pathogens (either “SARS-CoV-1-like Pathogen” or “SARS-CoV-2-like Pathogen”) and vaccination strategies (either ring-vaccination or spatially targeted vaccination campaigns).

Within this framework, the total number of secondary infections produced by individual *i, N_i_* is drawn from an offspring distribution as follows:

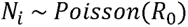

i.e. with the offspring distribution being a Poisson distribution with mean equal to the basic reproduction number of the pathogen, *R*_0_).

Each of these *N_i_* infections generated by individual 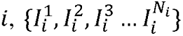 is then assigned a time of infection drawn from the generation time distribution *T_G_*, with this taking the form of:

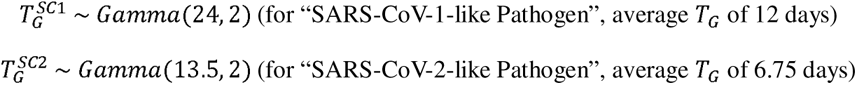

Each infection has an independent probability of being asymptomatic. Symptomatic infections develop symptoms following an incubation period, which is drawn from the incubation period distribution *T_I_*, with this taking the form of:

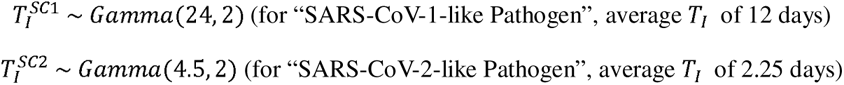

We note here that the expectation of the ratio 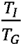 describes the average proportion of transmission that we expect to be presymptomatic i.e. the fraction of infections that are generated before the infector develops symptoms.

Each infection also has an independent probability of being hospitalised by the infection, with the time of hospitalisation relative to time of infection drawn from a time-to-hospitalisation distribution *T_H_*, with this taking the form of:

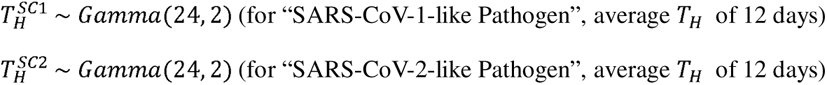

We used this framework to simulate the impact of two different vaccine-based containment strategies utilising the BPSV. These were ring-vaccination (where detection of symptomatic cases triggers reactive vaccination of all contacts of that case) and spatially targeted vaccination (where hospitalised cases trigger a vaccination campaign that seeks to vaccinate all individuals in a defined geographic area).

#### 1.2 Modelling Ring-Vaccination Campaigns

Within the ring-vaccination framework, following identification of the pathogen (assumed here to occur 21 days following a pathogen spillover event with 5 individuals initially infected), detection of a new symptomatic infection triggers reactive vaccination of all their contacts, which occurs after a delay of 2 days, reflecting the time to identify, notify and administer vaccination to contacts. Vaccinated individuals go on to develop immunological protection following vaccination at a delay of *d_V_* days. During this vaccination campaign, a number of outcomes are possible for the contacts of index cases that would otherwise be infected:

1. The individual is successfully vaccinated during the ring-vaccination campaign, protection arises before they would otherwise be infected, and their infection is successfully averted.
2. The individual is successfully vaccinated during the ring-vaccination campaign, protection arises before they would otherwise be infected, but this protection fails to avert their infection.
3. The individual is successfully vaccinated during the ring-vaccination campaign but infected before vaccine-derived protection arises.
4. The individual is infected before the ring-vaccination campaign can be carried out.
5. The individual is not identified by the ring-vaccination strategy and so is not vaccinated (assumed here to be 20% of contacts).

For individuals who receive vaccination and develop robust protection before they would otherwise be infected, whether or not they are still infected despite vaccination is given by a draw from a Bernoulli distribution with probability *p* = (1 - *V_I_*), where *V_I_* is the BPSV efficacy against infection. In individuals for whom protection has developed but who are infected anyway, we assume that BPSV vaccination renders breakthrough infections less infectious and hence these individuals have reduced transmissibility. The total number of secondary infections produced by vaccinated individual *i*, 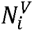 is drawn from the following modified offspring distribution:

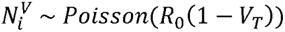

where *V_T_* is the BPSV efficacy against onwards transmission in vaccinated individuals with breakthrough infections. We note here that individuals with an asymptomatic infection are assumed to be undetected by the health-system and so do not trigger ring-vaccination.

#### 1.3 Modelling Spatially-Targeted Vaccination Strategies

To model the spatially targeted vaccination strategy, we modify the branching process such that each new infection is imbued with a set of geographical coordinates (their “home address”) {*x_i_, y_i_* }. The coordinates of these newly generated infections depend on the coordinates of the infector and a spatial kernel, which is a distribution describing the probability of the home addresses of two directly linked infections being separated by a certain amount of distance. Together with a direction, these factors determine the “home address” of newly generated infections. Specifically, if individual *i* is the infector with coordinates {*x_i_, y_i_*}, then the coordinates of infection *j* are generated using the following method:

− **Step 1:** Draw distance *d_i→j_* between infector *i* and infectee *j* from *d_i→j_* ∼ *NegativeBinomial*(*mu* = *µ*, *size* = 4)
− **Step 2:** Draw the direction of distance *d_i→j_* between infector *i* and infectee *j* from *Uniform*(0, 2π)

Simple trigonometric identities can be used to calculate the new coordinates {*x_j_, y_j_*} from an origin (here {*x_i_, y_i_*}), a distance and a direction. In practice however, the exact spatial kernel will depend on specific features of the setting, population and pathogen being considered. We therefore considered a simplified case whereby instead of explicitly specifying a spatial kernel in absolute terms, we describe it relative to the radius of the spatial vaccination campaign that is implemented.

Within this framework, the spatially targeted vaccination campaign is triggered by hospitalised infections. Hospitalised infections accumulate until a certain threshold is reached *T_H_*, after which the pathogen is said to have been “detected”. Following detection (and an assumed operational delay of 2 days), all eligible individuals within a certain spatial radius of the home address of the case which triggered the detection are vaccinated. During this vaccination campaign, a number of outcomes are possible for individuals:

1. The individual is successfully vaccinated and protection arises before they would otherwise be infected.
2. The individual is successfully vaccinated before they are infected but this protection fails to avert their infection
3. The individual is successfully vaccinated but infected before vaccine-derived protection arises.
4. the individual is infected before the vaccination campaign is carried out
5. The individual resides in an area outside the spatial radius of the vaccination campaign, is not vaccinated, and later becomes infected.
6. The individual chooses not to receive the vaccine (assumed here to be 20% of individuals).

Assumptions of BPSV properties and its impact on reduced transmissibility in breakthrough infections are the same as for the ring-vaccination strategy.

#### 1.4 Branching Process Vaccination Analyses

For both strategies, we simulate the proportion of outbreaks contained (defined as a final size of <10,000 infected individuals) and explore the prospects for containment whilst varying:

- **Pathogen Epidemiological Properties:** including the basic reproduction number, generation time distribution, extent of pre-symptomatic transmission, proportion of asymptomatic infections and proportion of infections that are hospitalised.
- **Intrinsic BPSV Properties:** including vaccine efficacy against infection and against onwards transmission (i.e. the degree of reduced transmissibility in breakthrough infections) as well as the delay between vaccination and protection developing.
- **Vaccination Campaign-Related Factors:** including the spatial kernel (namely the size of the vaccination campaign radius relative to the average distance between infections) and surveillance system sensitivity (determining the number of hospitalised cases triggering the spatial vaccination campaign).

In all instances, presented results are based on the proportion of outbreaks successfully contained across 100 stochastic simulations per parameter combination and vaccination strategy considered; and these results are compared to scenarios in which the BPSV is not available (i.e. there is no vaccination). Models were implemented in the programming language R and code required to reproduce the simulations presented in this work is available at https://github.com/mrc-ide/diseaseX_modelling

**Table S1:** Description of the key parameters varied during branching process analyses of ring and spatially targeted BPSV vaccination. Central value describes the fixed value used during sensitivity analysis of other parameters; range describes the set of parameter values explored during the sensitivity analysis for that particular parameter.

|  | Central Value | Sensitivity Analysis Range | Description | Notes |
| --- | --- | --- | --- | --- |
| **Pathogen Parameters** |  |  |  |  |
| $R_0$ | Varied | 1.25 – 2.5 | Basic reproduction number, describing the average number of secondary infections generated by an index infection. | |
| $T_G$ | • 12 days “SC1-like”<br>• 6.75 days “SC2-like” | See Notes on RHS | Generation time distribution, describing the distribution of infection times between index infections and secondary infections. | A series of sensitivity analyses were carried out varying $T_G$ for “SC2-like Pathogen” pathogen whilst keeping the incubation period $T_I$ – which in turn affects the proportion of presymptomatic transmission occurring. Varied between 15% and 70% presymptomatic transmission. |
| $T_I$ | • 12 days “SC1-like”<br>• 2.25 days “SC2-like” | See Notes on RHS | Incubation period distribution, describing the distribution of times between infection and symptom onset in non-asymptomatic individuals. | Selected to give central values of 0% presymptomatic transmission for “SC1-like” and 34% presymptomatic transmission for “SC2-like”. |
| **BPSV Parameters** |  |  |  |  |
| $V_T$ | 35% | 30%-90% | BPSV efficacy against onwards transmission i.e. the BPSV-mediated reduction in transmissibility in vaccinated individuals who become infected (breakthrough infections) | Efficacy against infection and efficacy against onwards transmission assumed to covary. Sensitivity analyses of efficacy involve varying these two parameters together. |
| $V_I$ | 35% | 30%-90% | BPSV efficacy against infection | |
| $d_V$ | Varied | 0 days – 28 days | Delay (in days) between BPSV vaccination and protection developing | Time between receiving vaccination and immunological protection developing. We assume here that the BPSV is delivered as a single dose regimen. |
| **Other Parameters** |  |  |  |  |
| $d_{i \rightarrow j}$ | 25x | 1x – 100x | Spatial kernel distribution describing the distribution of distances between pairs of directly linked infections. | Expressed as the distance relative to the radius of the spatial vaccination campaign area. |
| $T_H$ | 10 | 1 – 100 | The cumulative number of hospitalised infections required to trigger the spatial vaccination campaign. | |

### 2. Supplementary Methods: Dynamical Compartmental Modelling Framework

#### 2.1 Overview of Modelling Framework

We explored the potential utility of the BPSV in vaccination campaigns focussed on rapid mass-vaccination of priority groups following pathogen detection to support disease burden reduction and relaxation of societal restrictions imposed to control transmission (as has been the case with SARS-CoV-2 vaccination campaigns). To do this, we adapted a previously published dynamical model of SARS-CoV-2 transmission used to evaluate and explore the impact of SARS-CoV-2 transmission on COVID-19 mortality during the pandemic (*1*).

Briefly, the model is an age-stratified SEIRS (susceptible-exposed-infectious-recovered-susceptible) model that explicitly models the progression of COVID-19 disease severity, the transition through various levels of healthcare, and the introduction of vaccination campaigns. In terms of modelling disease progression and differential disease severity, the modelling framework explicitly includes elements of the clinical pathway differentiating individuals by disease severity within clinical settings (e.g. those requiring a basic hospital bed and limited oxygen consumption, as well as those requiring a protracted ICU stay and mechanical ventilation). Transmission between age-groups depends on age-based contact matrices, assuming a constant transmission rate per contact. The model includes an explicit latent period between an individual becoming infected and subsequently becoming infectious, as well as age-dependent probabilities of hospitalisation and disease severity. The model’s vaccination pathway incorporates both a delay in the development of protection following vaccination, as well as the possibility of waning of vaccine protection. Both of these are assumed to follow an Erlang distribution with a shape of two and a mean duration controlled by a model parameter that can be altered to reflect different assumptions around waning. Susceptible, latent, and recovered individuals can be vaccinated. Importantly, because latent individuals can be vaccinated, it is possible for latent individuals to develop vaccine derived protection before realising their infection, though in practice the size of this group relative to susceptible or recovered individuals is small and the effect of this is minor. Complete details of the model are given in (*16, 51, 52*) and we focus below on the major elaborations and alterations made to adapt the model to simulate a future, hypothetical SARS-X pandemic and deployment of both a BPSV and (later) a disease-specific vaccine against the pathogen.

##### 2.1.1 Modelling BPSV and Disease-Specific Vaccine Distribution

Following activation of BPSV stockpiles, eligible individuals (here considered to be all those aged 60+) are vaccinated with the BPSV at a constant rate and to a level of coverage determined by the size of the BPSV stockpile relative to the size of the eligible population. We assume here that the BPSV is delivered as a single dose regimen but note that the model is flexibly able to accommodate a wide variety of delays in the development of protection through the delayed protection mechanisms described in the paragraph above. In all scenarios considered, the BPSV is only delivered to the 60+ population – individuals below this age do not receive it. Following introduction of the disease-specific vaccine, elderly individuals at the greatest risk (again, those aged 60+) are prioritised to receive the disease-specific vaccine – and both elderly individuals who received the BPSV and those who did not (e.g. because the size of the stockpile precluded it) are vaccinated. Following complete vaccination of the elderly high-risk groups with the disease-specific vaccine, the disease specific vaccine is then rolled out and distributed to all other age-groups > 15 years.

##### 2.1.2. Modelling BPSV and Disease-Specific Vaccination Effectiveness

We model two distinct forms of vaccine efficacy: efficacy against infection and efficacy against severe disease (reducing risk of hospitalisation and death) in breakthrough infections (i.e. individuals who were vaccinated but where vaccination failed to prevent the infection occurring). We make the assumption that protection is partial, and that vaccine efficacy is the same for all individuals considered. For those protected by both vaccine-derived and infection-derived immunity, we assume that the most protective effect is dominant – though note given the timeframes over which we simulate (typically no longer than 18 months), we expect minimal waning of immunity to have occurred.

#### 2.2 Hypothetical SARS-X Pandemic Scenario Construction & Baseline Parameterisation

We use this modelling framework to explore the potential impact of BPSV availability on disease burden during a future hypothetical SARS-X pandemic. In this hypothetical scenario, the BPSV has been manufactured and stockpiled ahead of the pandemic, enabling rapid deployment following pathogen detection (which is the trigger for initiation of the BPSV vaccination campaign). We also explicitly model a pathogen-specific vaccine, which we assume to have a more favourable efficacy profile than the BPSV but that can only be developed following detection of the novel pathogen and sequencing of its genome. It is therefore only available after a significant delay. Our baseline scenario assumes the BPSV has 75% efficacy against severe disease and 35% efficacy against infection whilst a future disease-specific vaccine is assumed to have vaccine efficacy of 95% against severe disease and 55% against infection. In both instances, we assume there is minimal waning of vaccine-derived immunity over the timescale of the simulation period considered and we assume that it takes 7 days for development of immunological protection to occur following receipt of the vaccine. For the disease-specific vaccine we explore a development timeline of either 250 or 100 days (reflecting recent estimates from CEPI around realistic and ambitious vaccine development timelines (*13*)). In our simulations, pathogen spillover is followed by a period of undetected circulation in the community. Hospitalisations due to the infection lead to pathogen detection and identification. We assumed detection occurred when daily incidence of hospitalised individuals reached 5 hospitalisations per day. The time for this to occur following spillover was calculated using the stochastic branching-process based framework described above.

Following pathogen identification, development of the pathogen-specific vaccine starts, and after an assumed delay of 7 days (reflecting delays around decision making and activation of stockpiles), mass vaccination of the elderly population (here assumed to be those aged 60+) with the BPSV begins. The size of the BPSV stockpile is assumed sufficient to vaccinate 80% of the elderly population, with health systems capabilities able to vaccinate the population at a rate of 3.5% of the population per week, leading to completion of BPSV vaccination campaign within 3 weeks. All age-groups except those under 15 are eligible to receive the disease-specific vaccine, with rollout of this vaccine (sufficient to achieve a coverage of 80% of the population) occurring in the oldest age-groups first. We assume an R0 of 2.5 and an average generation time of 6.7 days, in-keeping with estimates derived for the original Wuhan-1 strain, as well as a severity profile and age-specific IFR similar to that of SARS-CoV-2 (*53*), adjusted to give an overall population-level IFR of 1%. We assume a demographic population age-structure matching the age-distribution of the World Bank Upper Middle-Income Country with the median age, and for the purposes of our scenarios assume no healthcare constraints that limit the ability of hospitalised individuals to access adequate medical care. We note that relaxing this assumption and imposing limited healthcare availability (and associated excess mortality risk arising from insufficient medical care) would only serve to increase our estimates of BPSV impact, and that previous analyses exploring the impact of COVID-19 vaccination on mortality have shown that direct protection was the main driver of deaths averted, with very few averted by the reduction in healthcare capacity required (*1*).

We explore 9 different scenarios varying the duration, stringency and timing of non-pharmaceutical interventions imposed in response to identification of the novel pathogen (shown in **Figure 2B**). Three stringency levels are considered: i) no NPIs (and so R is equal to R0); ii) a minimal and limited set of NPIs reducing transmission by 25%; and iii) a stringent set of NPIs sufficient to reduce Rt to 0.9. Whilst **Figure 2** considers 9 scenarios spanning a wide range of possible NPI responses, we use three central NPI scenarios for the rest of the analyses presented here. These are 1) “minimal NPI scenario” which assumes a short imposition of limited NPIs between pathogen identification and completion of the BPSV campaign; a 2) “moderate NPI scenario” involving imposition of the limited NPIs in response to pathogen identification. These then last in full until the end of BPSV campaign, whereafter they are gradually released and relaxed, and lift completely upon completion of the disease-specific vaccination campaign; and a 3) “stringent NPI scenario” which sees stringent NPIs implemented until the BPSV campaign is complete, followed by imposition of more limited NPIs which gradually lift until the disease-specific vaccination campaign is complete. We additionally carried out a series of sensitivity analyses varying a number of the model parameters described above. In all cases, we varied model parameters in a univariate manner whilst keeping all other parameter estimates identical to the baseline parameters described above. A detailed description of the exact parameters varied, and the parameter ranges used are described in **Table 1** and in the **Supplementary Table 2**.

For all scenarios, we calculated the deaths averted due to BPSV vaccination by subtracting the estimated SARS-X deaths from the simulation with both BPSV and disease-specific vaccines from the estimated number of SARS-X deaths under a scenario where only the disease-specific vaccine is available and report the number of deaths averted per 1,000 population. To calculate the NPI index featured in **Figure 2 of the main text** we construct a composite measure that considers both the duration and stringency of NPIs imposed in response to the hypothetical pandemic. This composite measure was calculated by first calculating the relative stringency of each set of NPIs, defined as the % reduction in the R0 by the NPIs (with a higher % reduction reflecting more stringent and costly NPIs). We then multiplied this stringency by the number of days spent under those NPIs to construct an index considering both stringency and duration of NPIs.

#### 2.3 Retrospective Evaluation of Potential Impact During SARS-CoV-2 Pandemic

Using previously published model fits calibrated within a Bayesian framework to excess mortality data (known to be a more complete measure of pandemic mortality, especially in LMIC settings with less robust vital registration) (*16*), we explored the potential impact that a stockpiled BPSV could have had on COVID-19 mortality in the first year of the pandemic. We used the resulting model fits to estimate the time-varying reproductive number, Rt, and its associated uncertainty by sampling 100 draws from the estimated posterior distribution of Rt from these previous fits. To estimate the impact of BPSV, we simulated a counterfactual scenario for each sampled Rt trajectory in which BPSV were introduced following globally reported COVID-19 deaths reached a certain threshold (varied between 1 and 1000 cumulative global deaths), under the assumption that all countries have access to a BPSV stockpile sufficient to vaccinate 60% of their eligible elderly population (here assumed to be those aged 60+) at a rate representing the average COVID-19 vaccination rate for each World Bank Income Strata based on data from Our World In Data (*32*); and under the strong assumption that availability of the BPSV would not have altered the NPIs imposed in response to SARS-CoV-2 (and hence alterations to Rt due to NPIs would be the same across both scenarios). We then calculated the deaths averted as a result of BPSV vaccination by subtracting the estimated COVID-19 deaths from the simulation with BPSV vaccines included from the estimated COVID-19 deaths from the simulation with only the real-world vaccination campaign included and reported the median deaths averted per 1,000 population.

**Table S2:**
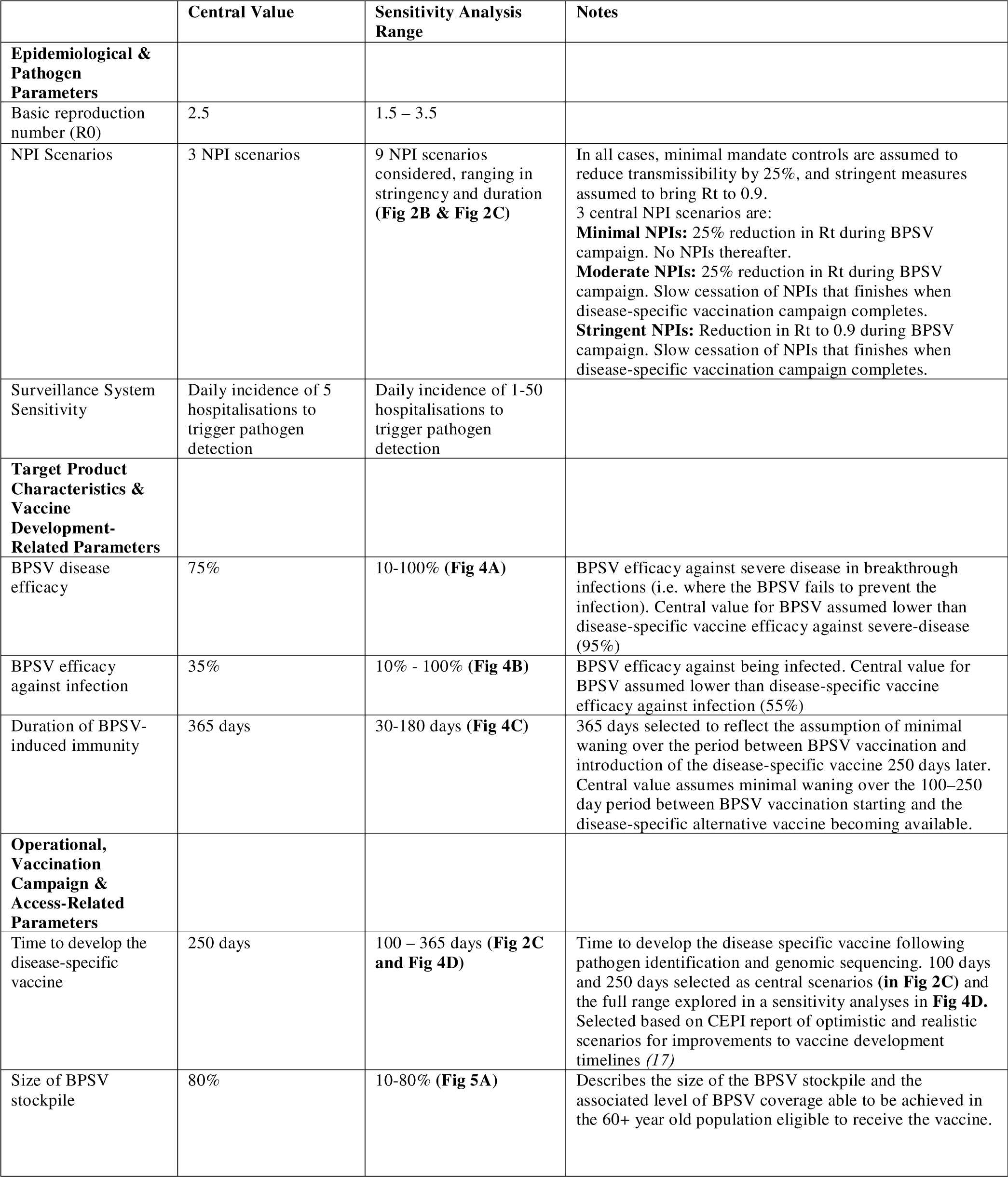
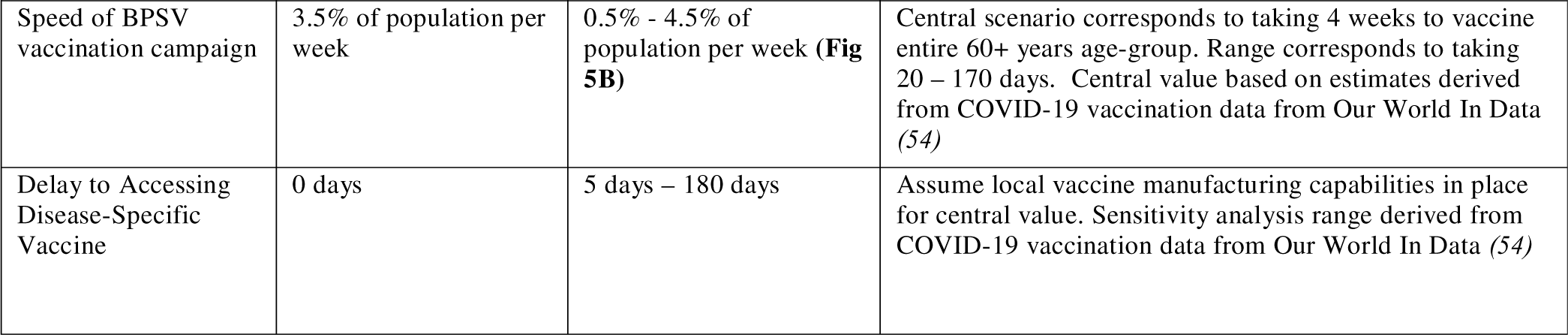
Description of model parameters varied in dynamical compartmental modelling of mass-vaccination of high-risk populations with BPSV. Central value describes the fixed value used during sensitivity analysis of other parameters; range describes the set of parameter values explored during the sensitivity analysis for that particular parameter. Parameter estimates were selected to replicate the approximate epidemiological properties of SARS-CoV-2, and complete set of model parameters used can be found in (*16, 51, 52*) and https://github.com/mrcide/diseaseX_modelling.

|  | Central Value | Sensitivity Analysis Range | Notes |
| --- | --- | --- | --- |
| **Epidemiological & Pathogen Parameters** |  |  |  |
| Basic reproduction number (R0) | 2.5 | 1.5 – 3.5 |  |
| NPI Scenarios | 3 NPI scenarios | 9 NPI scenarios considered, ranging in stringency and duration (**Fig 2B & Fig 2C**) | In all cases, minimal mandate controls are assumed to reduce transmissibility by 25%, and stringent measures assumed to bring Rt to 0.9.<br>3 central NPI scenarios are:<br>**Minimal NPIs:** 25% reduction in Rt during BPSV campaign. No NPIs thereafter.<br>**Moderate NPIs:** 25% reduction in Rt during BPSV campaign. Slow cessation of NPIs that finishes when disease-specific vaccination campaign completes.<br>**Stringent NPIs:** Reduction in Rt to 0.9 during BPSV campaign. Slow cessation of NPIs that finishes when disease-specific vaccination campaign completes. |
| Surveillance System Sensitivity | Daily incidence of 5 hospitalisations to trigger pathogen detection | Daily incidence of 1-50 hospitalisations to trigger pathogen detection |  |
| **Target Product Characteristics & Vaccine Development-Related Parameters** |  |  |  |
| BPSV disease efficacy | 75% | 10-100% (**Fig 4A**) | BPSV efficacy against severe disease in breakthrough infections (i.e. where the BPSV fails to prevent the infection). Central value for BPSV assumed lower than disease-specific vaccine efficacy against severe-disease (95%) |
| BPSV efficacy against infection | 35% | 10% - 100% (**Fig 4B**) | BPSV efficacy against being infected. Central value for BPSV assumed lower than disease-specific vaccine efficacy against infection (55%) |
| Duration of BPSV-induced immunity | 365 days | 30-180 days (**Fig 4C**) | 365 days selected to reflect the assumption of minimal waning over the period between BPSV vaccination and introduction of the disease-specific vaccine 250 days later. Central value assumes minimal waning over the 100–250 day period between BPSV vaccination starting and the disease-specific alternative vaccine becoming available. |
| **Operational, Vaccination Campaign & Access-Related Parameters** |  |  |  |
| Time to develop the disease-specific vaccine | 250 days | 100 – 365 days (**Fig 2C and Fig 4D**) | Time to develop the disease specific vaccine following pathogen identification and genomic sequencing. 100 days and 250 days selected as central scenarios (**in Fig 2C**) and the full range explored in a sensitivity analyses in **Fig 4D**. Selected based on CEPI report of optimistic and realistic scenarios for improvements to vaccine development timelines (17) |
| Size of BPSV stockpile | 80% | 10-80% (**Fig 5A**) | Describes the size of the BPSV stockpile and the associated level of BPSV coverage able to be achieved in the 60+ year old population eligible to receive the vaccine. |
| Speed of BPSV vaccination campaign | 3.5% of population per week | 0.5% - 4.5% of population per week (**Fig 5B**) | Central scenario corresponds to taking 4 weeks to vaccinate entire 60+ years age-group. Range corresponds to taking 20 – 170 days. Central value based on estimates derived from COVID-19 vaccination data from Our World In Data (54) |
| Delay to Accessing Disease-Specific Vaccine | 0 days | 5 days – 180 days | Assume local vaccine manufacturing capabilities in place for central value. Sensitivity analysis range derived from COVID-19 vaccination data from Our World In Data (54) |

**Figure S1:**
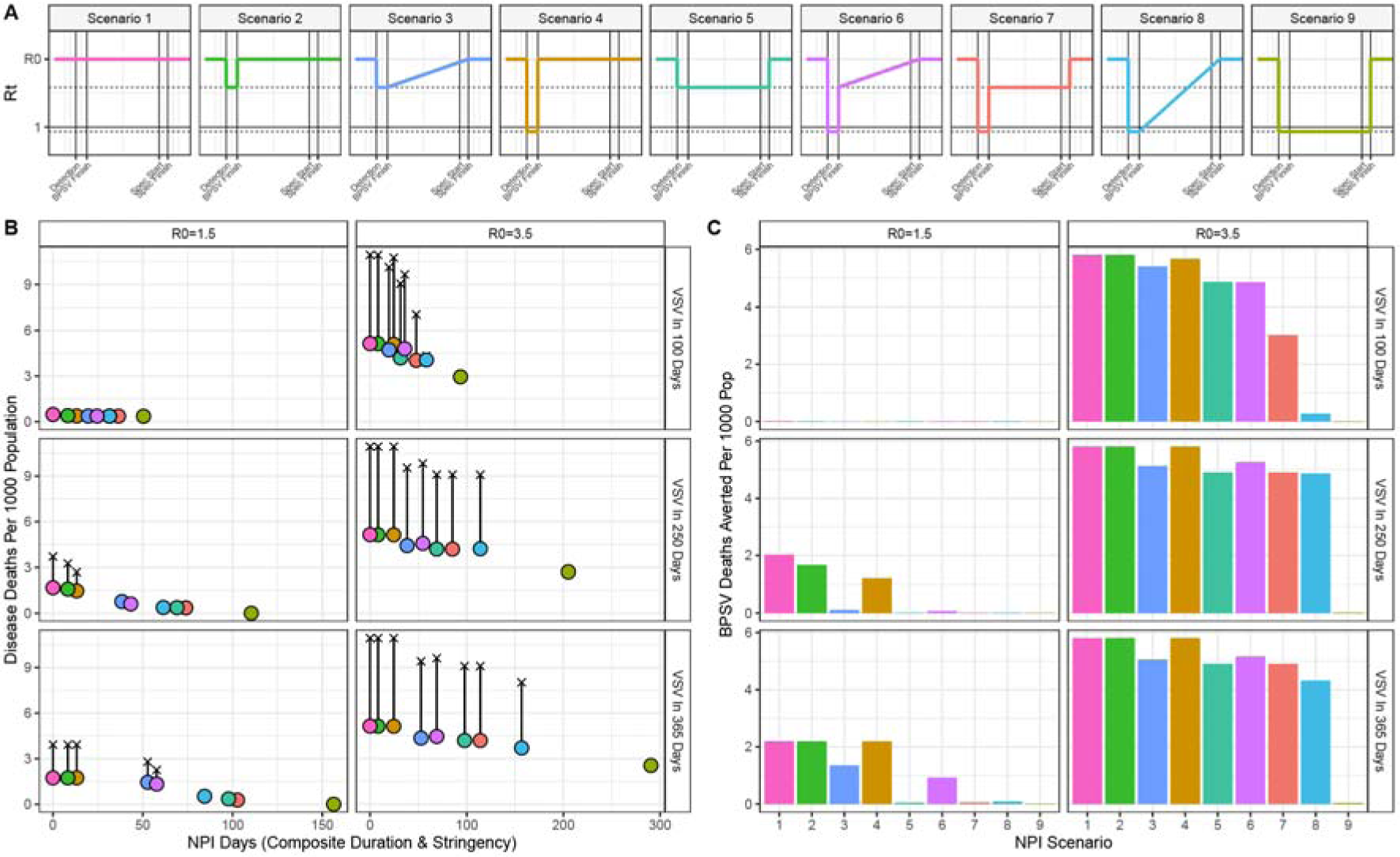
The influence of NPIs, R0 and VSV development timelines on BPSV impact during a SARS-X pandemic. **(A)** Time-varying reproduction number (Rt) profiles for the different non-pharmaceutical intervention (NPI) scenarios imposed in response to the epidemic that are considered for the analyses presented here. These Scenarios differ by assumed stringency (either no measures, a minimal mandate reducing transmission by 25% or stringent measures reducing Rt to 0.9), duration (either until the BPSV campaign is completed or the disease specific vaccination campaign is completed) and the nature by which these NPIs are relaxed (either instantaneous or gradual). **(B)** BPSV impact on disease burden for each NPI scenario, assuming the VSV is available 100 days (top-row), 250 days (middle row) or 365 days (bottom row) following detection, for an R0 of 1.5 (left hand column) or 3.5 (right hand column). Uncoloured crosses indicate scenario without BPSV (VSV only); points indicate scenarios where BPSV is available, coloured according to NPI scenario. **(C)** Deaths averted by the BPSV, coloured by NPI scenario and stratified by each unique VSV and R0 scenario considered here.

**Figure S2:**
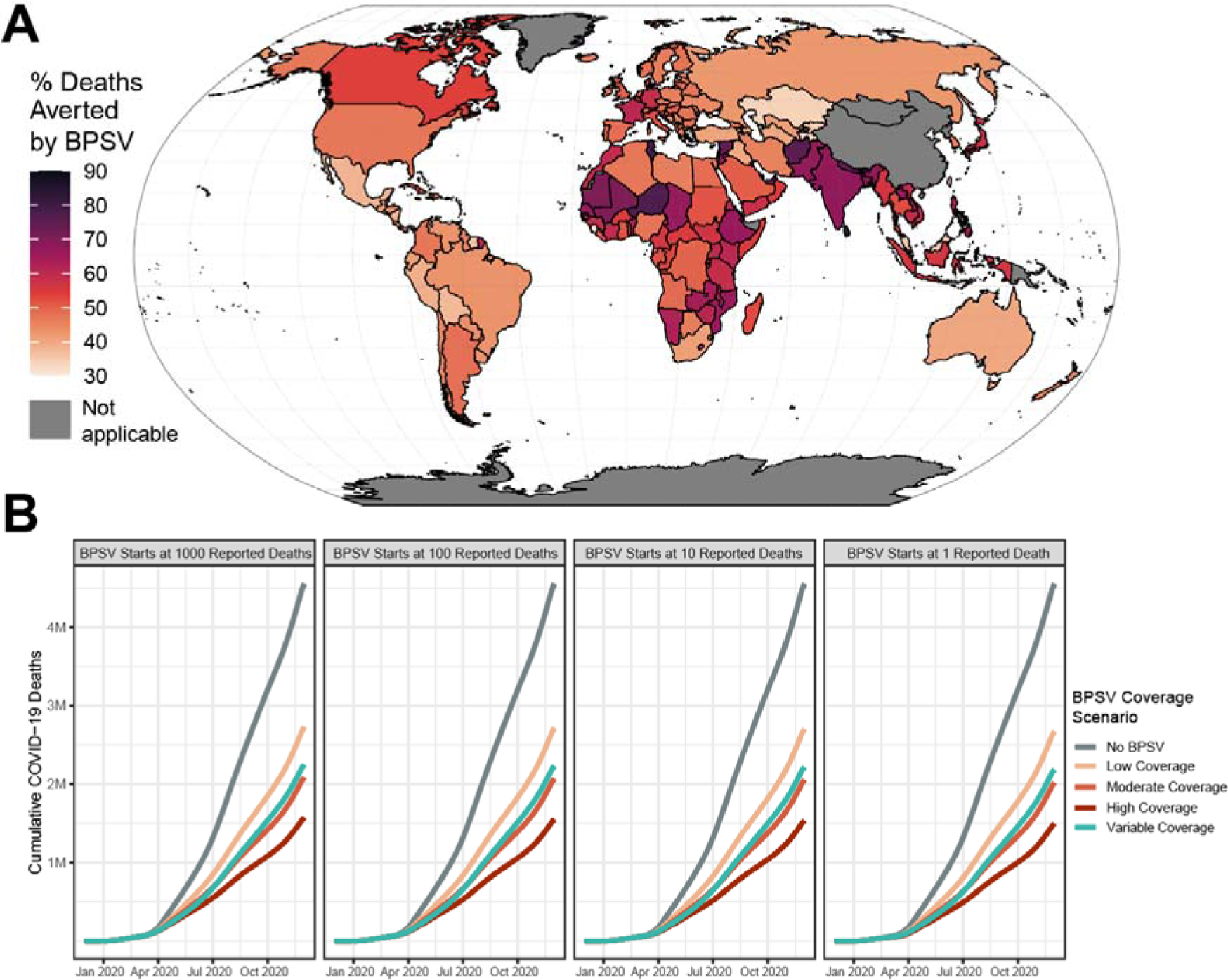
Assessing variation in BPSV deployment time and achieved coverage on impact. **(A)** Modelled impact of the BPSV during the first year of the COVID-19 pandemic in different countries around the world, assuming stockpile size sufficient to vaccinate 60% of each country’s eligible population (“Moderate coverage” scenario). Country colour indicates the percentage of COVID-19 deaths occurring in the first year of the pandemic that could have been averted if a BPSV had been available. **(B)** Cumulative global COVID-19 deaths during the first year of the pandemic without (grey) the BPSV, and with the BPSV (coloured lines). Low coverage = BPSV stockpile size sufficient to vaccinate 40% of elderly population; Moderate coverage = 60%; High coverage = 80%. Variable coverage indicates size of stockpile varies according to the World Bank Income Group each country belongs to (LIC = 20%, LMIC = 40%, UMIC = 60%, HIC = 80%). Facets indicate the number of globally reported COVID-19 deaths required for activation of BPSV stockpiles and initiation of eligible population vaccination.

**Figure S3:**
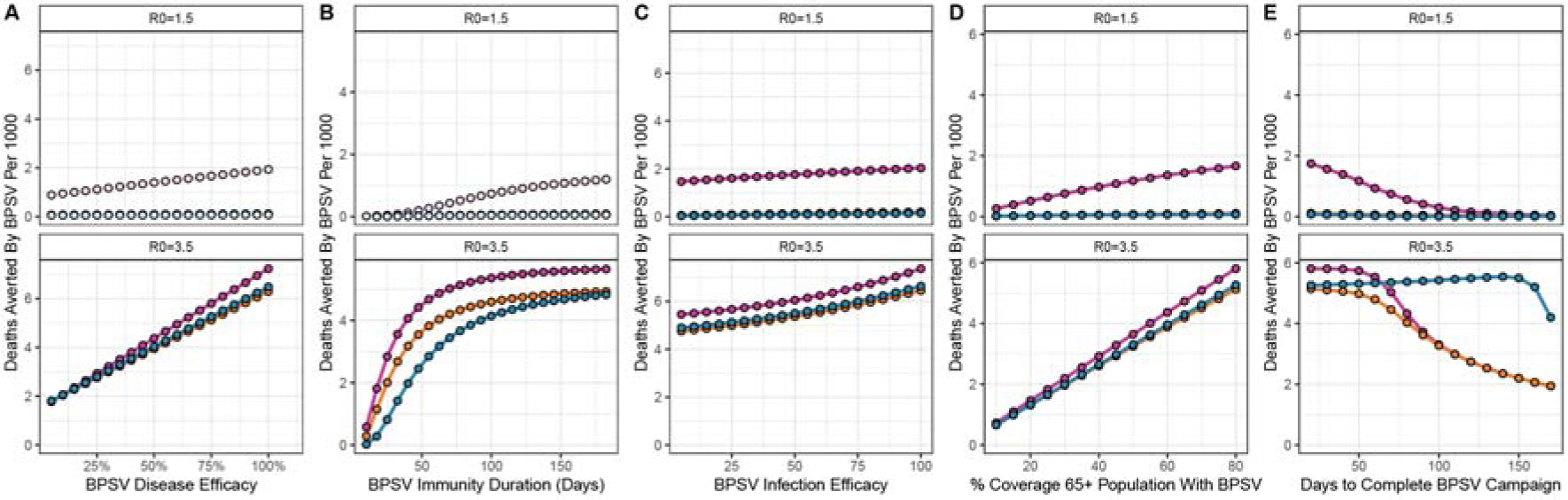
Dependency of BPSV impact on intrinsic vaccine properties, vaccination campaign dynamics and virus properties. Sensitivity analyses exploring the sensitivity of BPSV impact to intrinsic BPSV properties and factors governing the speed, availability and coverage of the BPSV vaccination campaign, as well as the basic reproduction number of the virus. **(A)** Deaths averted by the BPSV (per 1,000 population) and BPSV efficacy against severe disease. Results coloured according to NPI scenario considered (pink = minimal, orange = moderate, blue = stringent), for R0=1.5 (top row) and R0=3.5 (bottom row). **(B)** As for (A) but for BPSV efficacy against infection. **(C)** As for (A) but for BPSV immunity duration. **(D)** As for (A) but for BPSV stockpile size (and associated coverage of the target population that can be achieved). **(E)** As for **(A)**, but for the rate of vaccination during the BPSV campaign (and the associated time taken to vaccinate all eligible and willing individuals).

